# Population-based prevalence surveys during the COVID-19 pandemic: a systematic review

**DOI:** 10.1101/2020.10.20.20216259

**Authors:** Vinícius Bonetti Franceschi, Andressa Schneiders Santos, Andressa Barreto Glaeser, Janini Cristina Paiz, Gabriel Dickin Caldana, Carem Luana Machado Lessa, Amanda de Menezes Mayer, Julia Gonçalves Küchle, Paulo Ricardo Gazzola Zen, Alvaro Vigo, Ana Trindade Winck, Liane Nanci Rotta, Claudia Elizabeth Thompson

**Affiliations:** Center of Biotechnology, Graduate Program in Cell and Molecular Biology (PPGBCM), Universidade Federal do Rio Grande do Sul (UFRGS), Porto Alegre, RS, Brazil; Undergraduate Program in Biomedicine, Universidade Federal de Ciências da Saúde de Porto Alegre (UFCSPA), Porto Alegre, RS, Brazil; Graduate Program in Pathology, Universidade Federal de Ciências da Saúde de Porto Alegre (UFCSPA), Porto Alegre, RS, Brazil; Graduate Program in Epidemiology, Universidade Federal do Rio Grande do Sul (UFRGS), Porto Alegre, RS, Brazil; Graduate Program in Health Sciences, Universidade Federal de Ciências da Saúde de Porto Alegre (UFCSPA), Porto Alegre, RS, Brazil; Undergraduate Program in Biomedical Informatics, Universidade Federal de Ciências da Saúde de Porto Alegre (UFCSPA), Porto Alegre, RS, Brazil; Department of Internal Medicine, Universidade Federal de Ciências da Saúde de Porto Alegre (UFCSPA), Porto Alegre, RS, Brazil; Department of Statistics, Universidade Federal do Rio Grande do Sul (UFRGS), Porto Alegre, RS, Brazil; Department of Exact and Social Applied Sciences, Universidade Federal de Ciências da Saúde de Porto Alegre (UFCSPA), Porto Alegre, RS, Brazil; Department of Diagnostic Methods, Universidade Federal de Ciências da Saúde de Porto Alegre; Department of Pharmacosciences, Universidade Federal de Ciências da Saúde de Porto Alegre (UFCSPA), Porto Alegre, RS, Brazil

**Keywords:** COVID-19, Severe acute respiratory syndrome coronavirus 2, Cross-Sectional Studies, Prevalence, Epidemiology, Infectious Diseases

## Abstract

Population-based prevalence surveys of COVID-19 contribute to establish the burden and epidemiology of infection, the role of asymptomatic and mild infections in transmission, and allow more precise decisions about reopen policies. We performed a systematic review to evaluate qualitative aspects of these studies, their reliability, and biases. The available data described 37 surveys from 19 countries, mostly from Europe and America and using antibody testing. They reached highly heterogeneous sample sizes and prevalence estimates. Disproportional prevalence was observed in minority communities. Important risk of bias was detected in four domains: sample size, data analysis with sufficient coverage, measurements in standard way, and response rate. The correspondence analysis showed few consistent patterns for high risk of bias. Intermediate risk of bias was related to American and European studies, blood samples and prevalence >1%. Low risk of bias was related to Asian studies, RT-PCR tests and prevalence <1%.

**One sentence summary:** Population-based prevalence surveys of COVID-19 until September 2020 were mostly conducted in Europe and Americas, used antibody testing, and had important risks of bias.

## Introduction

In December 2019, the third most important coronavirus in the twenty-first century (SARS-CoV-2) was identified as the causative agent of severe acute respiratory syndrome outbreak in Wuhan, Hubei province, China (*1,2*). The illness named COVID-19 spread rapidly around the world, acquiring pandemic status on March 11, 2020 (*3*). As of 14 October, 2020, there are ∼38 million confirmed cases and ∼1.1 million reported deaths in 216 countries, areas or territories. More than 50% of these cases were reported in the USA, India and Brazil, the worst-hit countries (*4,5*).

According to the current evidence, the main form of SARS-CoV-2 spreading is through human-to-human transmission via respiratory droplets and contact routes (*6*). The standard diagnostic testing method is the Real-Time reverse transcription-PCR (RT-PCR) test (*7,8*), which is able to detect current infections and it is recommended for people with COVID-19 symptoms and for all close contacts of the confirmed cases. A complementary approach is to use antibody tests (e. g. point-of-care test or enzyme linked immunosorbent assay) to detect a past infection and the production of antibodies (IgM and/or IgG) against SARS-CoV-2 (*8*).

COVID-19 causes diverse degrees of illness, ranging from asymptomatic infection to severe pneumonia (*9*). However, surveillance is only based on the confirmed cases, which can represent an underestimation of total cases due to non-testing in mildly affected or asymptomatic individuals. Population-based prevalence surveys can help to establish the disease epidemiology, the burden of infection, the role that asymptomatic and mild infections play in the transmission and to enable precise evidence-based decisions about control and reopen policies while no pharmacological intervention is available (*10*). Moreover, accurate estimates of the basic reproduction number, of exposed and susceptible populations, and the fatality rates can be obtained (*11,12*).

Statistical extrapolations will only be reliable for the population if (i) the sample of individuals is sufficient, random, and representative of the general population, (ii) if the measurements are standardized, and (iii) if the tests used have adequate sensitivity and specificity, among other factors (*13*). For example, a recent systematic review and meta-analysis evaluated the diagnostic accuracy of serological tests in 40 studies. The conclusion indicated that the use of existing point-of-care serological tests is not supported by available evidence due to low performance (*14*). Thus, a critical evaluation of these parameters is necessary to verify the reliability of the population-based surveys of COVID-19.

We performed a systematic review to evaluate and summarize the main results regarding the COVID-19 prevalence obtained through population-based surveys, their reliability and biases. Our main aims were to evaluate the qualitative aspects of these studies and to compile methodology practices that can influence positively or negatively the prevalence estimates.

## Methods

### Registration and Reporting

The protocol for this systematic review was registered on PROSPERO (<u>CRD42020202186</u>) and it is available in full on Appendix 1. Reporting was conducted according to Preferred Reporting Items for Systematic Reviews and Meta-Analyses (PRISMA) (Appendix 2, Checklist).

### Search Strategy

Systematic literature searches for published and unpublished (preprint) articles were conducted from 15 July to 05 September, 2020. MEDLINE (accessed via PubMed), Excerpta Medica dataBASE (EMBASE), bioRxiv, and medRxiv databases were searched using the following controlled vocabulary heading and terms: “seroprevalence”, “prevalence”, “serology”, “immunoassay”, “enzyme linked immunosorbent assay”, “real time polymerase chain reaction”, “cross-sectional study”, “population screening”, “severe acute respiratory syndrome coronavirus 2”, “COVID-19”. These terms and their synonyms were combined using logical operators and adapted according to the searched database. Only articles published in English were retrieved. The complete search strategy for each database is on Appendix 2, Table 1.

### Inclusion and Exclusion Criteria

The review included cross-sectional or repeated cross-sectional studies using molecular or serological tests to estimate the prevalence of COVID-19 in municipalities, regions, states or countries around the world. Studies were excluded based on the following criteria: (i) non-cross-sectional studies, (ii) studies with correlation between COVID-19 and other diseases or health determinants, (iii) non-random selection of participants (e. g. convenience sampling), (iv) inclusion of a specific group of participants only (e. g., with comorbidities, pregnant, elderly, healthcare workers, pediatric patients), and (v) non-human samples.

### Article Screening and Data Extraction

Four pairs of authors (AMM and CLML, ABG and JGK, ASS and VBF, GDC and JCP) independently reviewed the titles and abstracts, in parallel, and included publications identified by either author for full-text review. These authors also reviewed full texts to determine which publications met the inclusion criteria and then re-analyzed the texts and supplemental materials to extract the following relevant information, when available: (i) authors, (ii) study location, (iii) coverage, (iv) study type, (v) random sampling method, (vi) period of testing, (vii) number of tests, (viii) biological samples, (ix) type of test used, (x) if test validation was performed, (xi) the test sensitivity and specificity, (xii) prevalence, and (xiii) statistical methods (Table). Disagreements in the screening and data extraction were discussed among the reviewers and, if consensus cannot be reached, a third reviewer (ATW) made the ultimate decision.

### Survey Quality

We assessed each survey quality by using the Joanna Briggs Institute (JBI) Critical Appraisal Checklist for Prevalence Studies (*13*). This tool evaluates nine domains: (D1) sample frame adequacy, (D2) recruitment method, (D3) sample size, (D4) study subjects and the setting, (D5) coverage, (D6) diagnostic methods, (D7) the reliability and standardization of measurements, (D8) statistical analysis, and (D9) the response rate. For each study, “yes”, “no” and “unclear” options were selected, meaning “low”, “high” and “unclear” risk of bias, respectively. The number of “yes” answers to these nine domains was counted, with a higher number of yes representing less risk of bias. Graphs considering each risk of bias domain across all studies were prepared using the robvis R package v. 0.3.0.900 (*15*).

### Definitions

Additional objective criteria were adopted for the survey quality assessment. For D4, the prevalence estimates should be stratified by conventional sex and age classes minimally. For D5, “no” was chosen when there was a lack of a subgroup representativity. If the response rate >70% or <70% with adequate sample size, “yes” was chosen. The option “unclear” was selected only if there was no information about the response rate in the article. For D6, a method was considered valid if the sensitivity >70%. For D7, self-sampling was considered as a practice of high risk of bias. In the case of a collection described by health professionals or trained individuals and using standardized methods, we assumed a low risk of bias. For D8, a minimum description of statistical methods was sufficient to classify the study as low risk of bias. For D9, if the response rate <70% without stratification or statistical management, the study was considered to have a high risk of bias. Response rate >70% or appropriate management of low response rate were related to a low risk of bias, while missing information about the proportion of tested in relation to the recruited individuals was associated with unclear.

### Data analysis

A correspondence analysis was performed to visualize in a low-dimensional graphic the relationships among categories of the row and column variables. The row variables (respective categories) were: (i) study continent (Africa, Asia, Europe, North America, and South America); (ii) coverage (country, region, and municipality); (iii) biological samples (uninformed, blood only, both swab and blood, serum/plasma, and swab only), (iv) test validation (external, uninformed, yes [internal], and RT-PCR [N/A: gold-standard]), (v) test sensitivity (<80%, 80-90%, 90-100%, unavailable, and RT-PCR [N/A: gold-standard]). The column variables (respective categories) were: (vi) prevalence (<1%, 1-3%, 3-5%, 5-20%, and >20%), and (vii) risk of bias (low [≤ 1 high risk], intermediate [1 < high risk ≤ 3], and high [> 3 high risk]) (see Survey Quality). Two studies (*18,34*) were split due to widely divergent prevalences reported in each part of the municipalities investigated. Therefore, despite the 37 studies included, 39 records were considered in this analysis. The PROC CORRESP from SAS Studio (Release 3.8, Enterprise Edition) available on the SAS OnDemand for Academics platform was used to perform the correspondence analysis.

## Results

Of 49 full-text articles screened, we excluded 12 (Appendix 2, Table 2), and identified 37 eligible for extensive review (Figure 1, Table). Of these, 23 (62.2%) were preprint, while 14 (37.8%) were peer-reviewed and published. Fifteen articles (40.5%) were from Europe, 8 (21.6%) from North America, 8 (21.6%) from South America, 5 (13.5%) from Asia, and 1 (2.7%) from Africa. The countries with the vast majority of population-based prevalence study initiatives were the United States (n=8; 21.6%), Brazil (n=7; 18.9%), and the United Kingdom (n=3; 8.1%). Importantly, 15 of the 16 studies in the Americas were conducted in the United States or Brazil, which are included in the TOP 3 of confirmed cases and deaths worldwide. In total, 19 countries had studies included in this analysis (Figure 2). Considering the coverage of these studies, 16 (43.2%) had regional (state / province / county) scope, 13 (35.1%) were restricted to municipalities, and 8 (21.6%) were nationwide studies.

**Figure 1.**
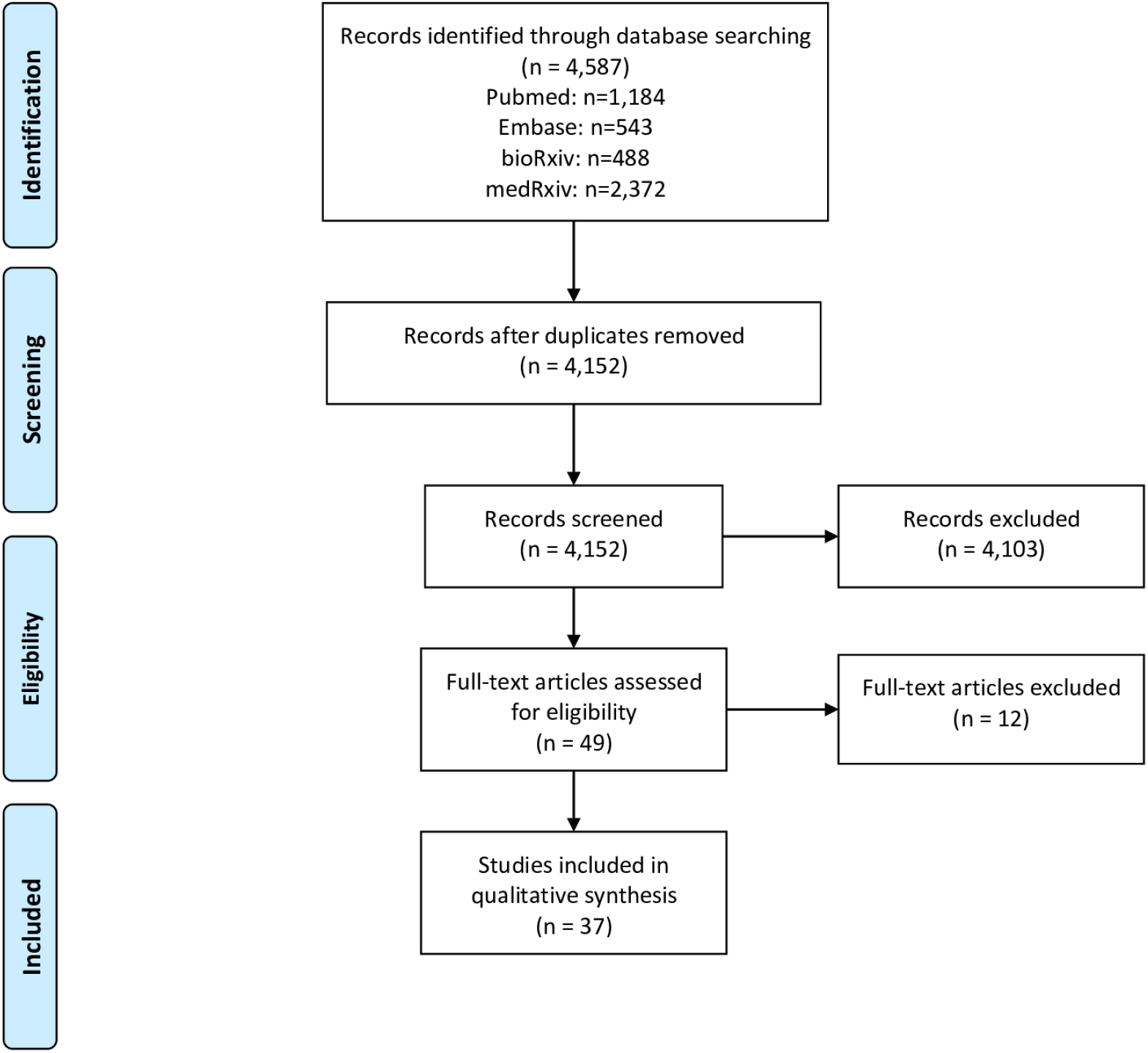
PRISMA Flowchart of the literature search.

**Figure 2.**
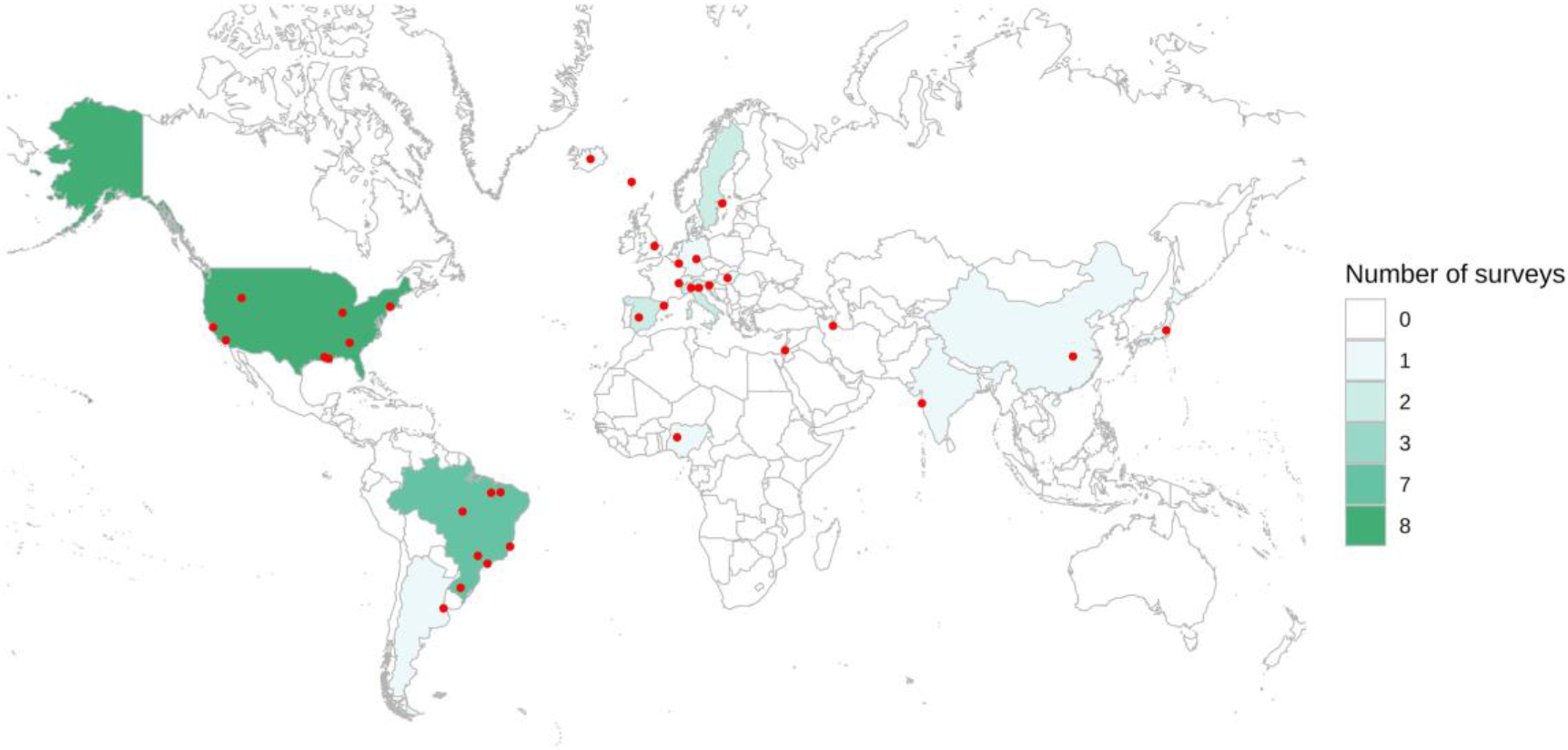
Map of countries and specific regions with prevalence surveys. Red dots represent regions and cities where the initiatives were performed. In nationwide studies, the point was placed in the center of the country.

The vast majority of studies (n=25; 67.6%) reported only antibody testing, while the exclusive use of RT-PCR were presented in 5 (13.5%), and both tests were conducted in 7 (18.9%) studies. The authors of 15 (46.9%) of the 32 studies that used serological tests reported their own validation test performance, while in 13 (40.6%) the validation performed by other studies or by the manufacturer were described. Excluding Wuhan’s (China) screening program that tested 9,899,828, at least 394,090 individuals were tested in the other 36 studies that reported the number of tests. However, this number was highly variable among studies (mean: 10,946.94, median: 1,990, standard deviation (SD): 27,382.34). Considering the periods of these surveys, most of them were conducted between April and July, 2020 (Figure 3).

**Figure 3.**
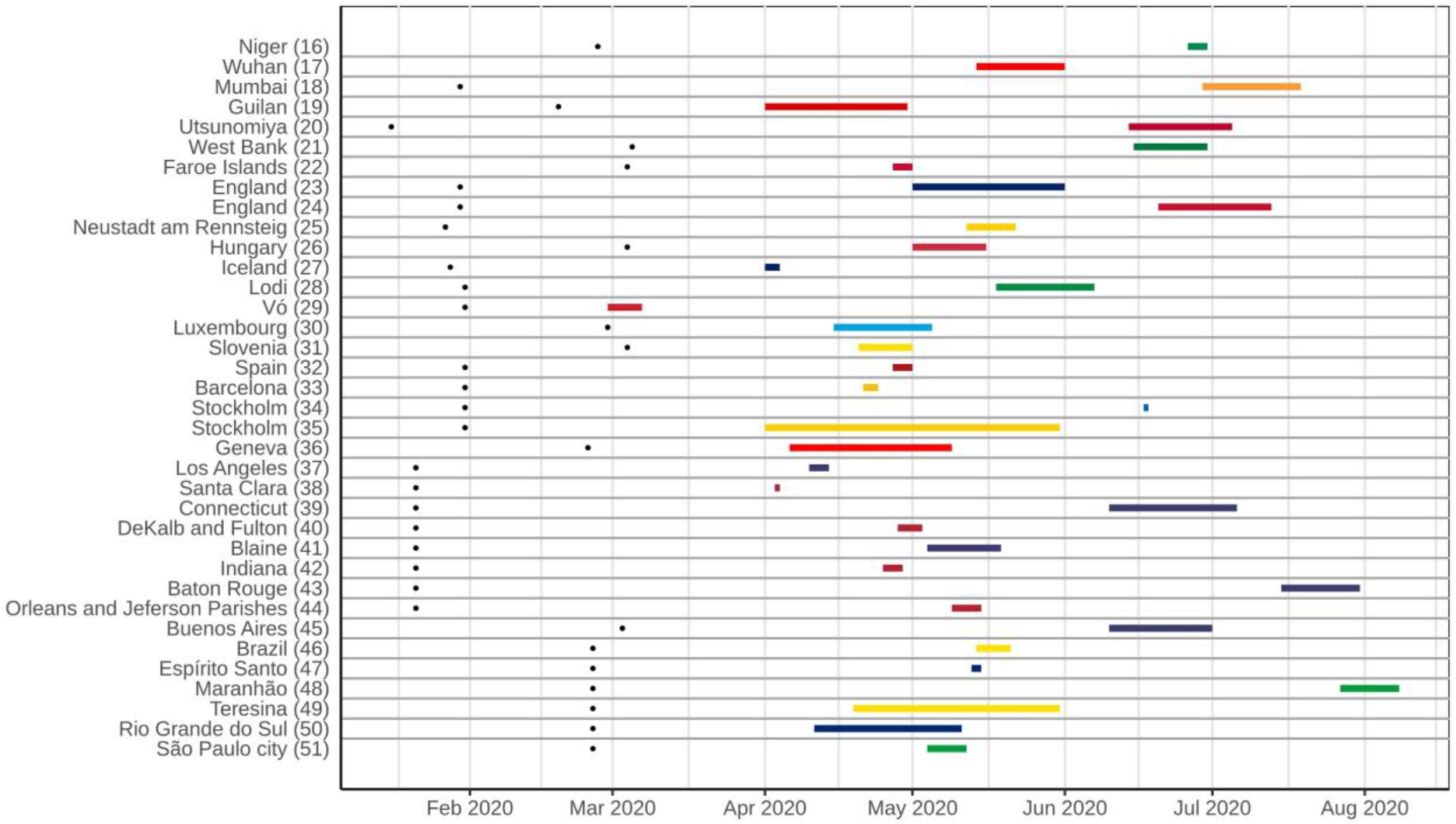
Timeline of population-based COVID-19 prevalence surveys conducted worldwide, with the duration of each survey and an overview of the most represented periods. Black dots on the left represent the date of the first confirmed case in the country of each survey.

Most studies (n=35; 94.6%) presented low risk of bias overall, but only one had low risk of bias in all nine domains (*32*). Two studies showed overall unclear risk of bias (*34,49*). Apart from these, another three studies had a sum of high and unclear risk of bias higher than the low risk of bias (*16,20,27*) (Appendix 2, Figure). Considering the nine domains established and three possible answers (low, unclear, and high), on average 6.35, 1.43 and 1.19 of each option was chosen, respectively. The median values were 6.0, 1.0 and 1.0 while the SDs were 1.44, 1.26 and 1.08. Considering the sum of results with some risk of bias (unclear and high), the mean, median and SD were 2.62, 3.0 and 1.46, respectively. Considering each domain in all studies, >75% low risk of bias across the studies were observed in five domains. On the other hand, three criteria (data analysis with sufficient coverage, measurements in standard way, and response rate adequacy) were adequate in <50% of the studies. The remaining domain (sample size) was adequate in ∼70% of the studies (Figure 4).

**Figure 4.**
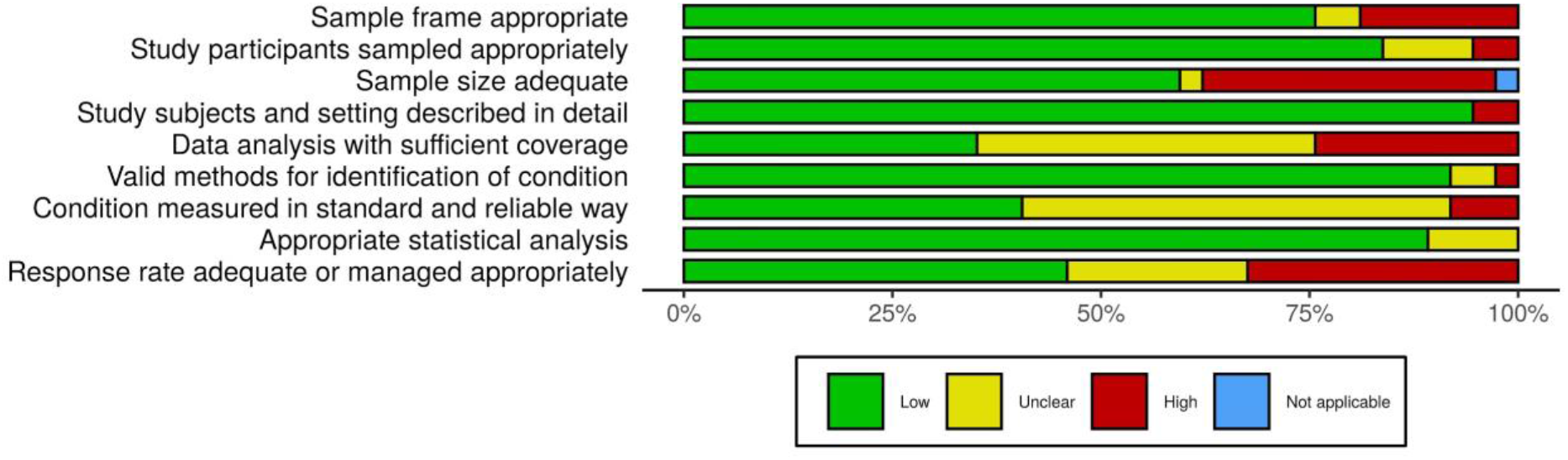
Risk of bias assessment summary table across all studies. *No weights were applied for different studies. †Not applicable was selected in “sample size adequate” because the study had zero prevalence (impossible to calculate the sample size required).

Considering the analysis of correspondence performed (Figure 5) among seven main variables (continent, coverage, biological samples, test validation, sensitivity, prevalence, and risk of bias), we found some important correlations. European, North- and South-American studies presented, in general, an intermediate risk of bias, while Asian studies tended to a low risk of bias. Regarding the coverage of the studies, both regional, nationwide and municipal studies presented an association with intermediate risk of bias.

**Figure 5.**
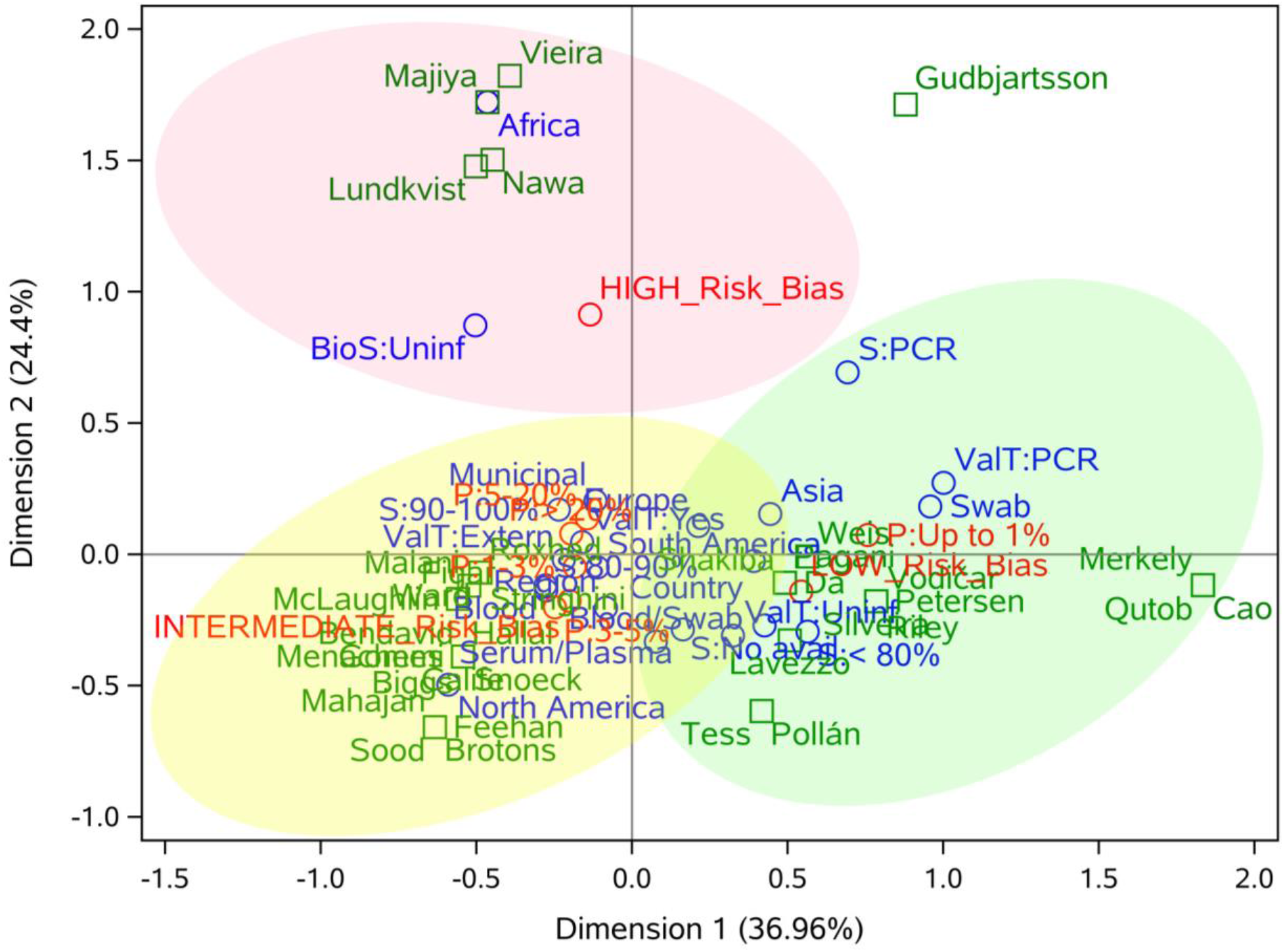
Correspondence analysis of seven important variables of population-based COVID-19 prevalence surveys. The authors of the studies were included as a supplementary variable and are represented in green, the categories of row (continent, coverage, biological samples, test validation, and sensitivity) and column (risk of bias and prevalence) variables are represented in blue and red, respectively. Light red, yellow, and green ellipses represent high, intermediate and low risk of bias, respectively. * BioS: Biological sample; S: sensitivity; P: prevalence; ValT: test validation.

Studies that performed molecular tests on naso- and oropharyngeal swabs (NPS) tended to have a low risk of bias, while those with blood samples were related to intermediate risk. Regarding prevalence, the majority of the studies with swab samples (RT-PCR) showed prevalence (P) <1%, while studies using only blood, or swab and blood, exhibited P>1%. Validation in serological tests had no significant impact on the quality of studies, since both external and internal validation were related to intermediate risk of bias. On the other hand, the use of the gold standard (RT-PCR) was associated with low risk of bias. P<1% was more frequent in studies with low risk of bias, while P>1% was associated with intermediate risk of bias.

## Discussion

We observed that important limitations of the studies were the low sample size and the low response rate (Figure 4). These factors influence heavily on reliable prevalence estimates (*53*). Moreover, the recruitment by letter, by mail or online may play a significant role in reducing the response rate and inadequately address the target population (*54,55*).

For example, in the Icelandic study (*27*), the authors discussed the small variation in the prevalence estimates between open invitation and random selection recruitments. However, the random selection methods were not detailed and the sample size to detect the estimated prevalence was not adequate (<2,529 individuals) (*56*). In the Slovenian study (*31*), despite being considered nationwide, the sample size was 1,366, which represented ∼7x less than necessary (10,179) (*56*) and there was no management of the low response rate (<50%). Some authors seem to have not been concerned with managing this issue because even though the response rate was low, there was still an adequate sample (*23,35,36,40,42*). Repeated cross-sectional studies featured a widely distinct prevalence estimate on each round (*29,36,49,50*). This trend might be caused by the ascending curve of infected people, following the epidemic’s natural course. Therefore, there was a need for different sample sizes for each period. Unfortunately, some studies did not yield adequate sample size in all rounds (*49, 50*).

The same proportion of studies validated their methods internally to report accuracy (*16,21,23,24,30,32,34-39,45,50,52*) or used sensitivity and specificity given by manufacturers or other external studies (*18-20,22,36,40-42,44-51*). We also noticed that it is quite unclear if the field teams followed standardized protocols for the data collection and testing. The absence of complete information resulted in a loss of quality in the methodological analysis (*13*), and we speculate that one rationale would be the editing process of these articles, which were published as letters or comments. In some studies (*23,24,35*), the samples were collected by the participants themselves, which causes an increase in the number of discarded samples and can reduce the sensitivity, especially of RT-PCR, highly influenced by a well done sample collection procedure (*57*).

However, the studies presented several strengths to highlight. Valid methods consistently stated the identification of the condition and the manufacturer indicated this accuracy. The majority of the sampling methods was conducted appropriately regarding randomness, and the participants were well described and stratified, thus mitigating possible selection bias (*18,19,24,26,28,30,33,37,39,43,44,46,48-51*). A strong trend was observed in relation to the sampling procedures used in Brazil. All studies used a standardized household sampling method based on census tracts (*46-48,50-52*) or healthcare units (*49*).

An interesting method of sample selection was the use of social networks ads targeting individuals by demographic and geographic characteristics and stratification, which despite being convenient inserts the biases of technology usage and the participation of people most likely to be infected. However, in these cases, statistical management seems to have been adequate to accommodate the sampling issues in the prevalence estimates (*38,43*). Biases were also introduced when volunteers were recruited, but data analysis was conducted properly in these cases (*41,43*). Nevertheless, these practices cover up important methodological issues despite minimizing the biases of studies and they should be avoided.

COVID-19 has an extensive spectrum of manifestation, including asymptomatic infection, mild disease, severe pneumonia, and death (*2,9*). Asymptomatic individuals may play an important role in disease transmission (*10*). The prevalence of asymptomatic infection in the community is still unclear, but essential to estimate the real COVID-19 prevalence. Infection rates rely on testing of symptomatic persons and may underestimate infection rates (*10,11,12*), which can be circumvented by surveying randomly recruited populations (*11*).

In fact, the asymptomatic rate of infection is quite hard to estimate. Nevertheless, we can consider some relevant observations. The proportion of symptom-free SARS-CoV-2 infection in most studies is higher than Severe Acute Respiratory Syndrome (SARS) (*58*) and Middle East Respiratory Syndrome (MERS) (*59*) coronaviruses epidemics, which was reported to lay between 0% and 7.5%. However, in the case of COVID-19, these rates were widely variable among PCR-positive and/or seropositive, ranging from 19.6% (*47*) to 69% (*23*). Older age groups were correlated with SARS-CoV-2 infection among symptomatic participants (*28,29*), and there was no statistical difference in the viral load of symptomatic versus asymptomatic (*29*). On the other hand, PCR- and antibody-negative participants also reported symptoms (*25,27*), raising the possibility of infection by other respiratory etiological agents (*27*). However, the comparability of asymptomatic rate estimates is hindered by different approaches applied, since the period of symptoms screening before the sampling ranged from one week (*23*) to several months (*26*).

Some studies demonstrated a disproportionate seroprevalence in black communities (*24,40,42-44,46,47,51*), multiracial, hispanic, indigenous, and asian persons (*24,38,39,43,44,46*), as well as in public-facing workers (*17,24,43*), and slums population (*18,45*). These data show the disparities that minority communities face to access healthcare systems, arisen from a complex relationship of social, environmental, economic, and structural inequities (*60,61*). Therefore, a priori knowledge of these trends in seroprevalence is essential for the sample design and for the instruction of field teams regarding protective measures in these surveys.

In the study from Stockholm (*34*), it was observed a significant difference in seroprevalence between the two areas (4.1% in middle-high income and 30% in lower income suburb). The authors related this high prevalence with cramped accommodation, which enhances cluster transmission, and with a majority of public-facing workers in the suburb. In Mumbai (*18*), the authors found a higher seroprevalence in the slums (54.1%) compared with non-slums (16.1%). Thus, it is discussed that the epidemic may be in advanced stages in slums due to higher population density.

The data from Brazilian studies (*46-52*) suggest that pandemic were highly heterogeneous in the country, with rapid growth in North and Northeast regions, and slow progression in the South and Center-West regions. These data demonstrate the impact of differences in demographics, urban infrastructure and income on the infection spreading and seroprevalence, emphasizing health inequality (*62,63*).

It is important to note that the data presented here are based on the articles until September 5, 2020. Therefore, more recent articles are not included in the analysis. In addition, previous pre-print articles can be currently published. In general, we believe that the peer review process should contribute to increase the quality of these currently unpublished articles with a higher risk of bias.

We have decided not to conduct a meta-analysis because of the prevalence heterogeneity among studies and the different stages of pandemic faced in the countries and continents at the time of each survey. Thus, a summary measure of meta-analysis would not be able to generalize overall findings sufficiently. In contrast, we found that a correspondence analysis was more able to detect the correlation among variables.

In this analysis, few consistent patterns were observed for studies with a high risk of bias, indicating that particular methodological choices of each study may affect its quality, not choices that are being made in many studies worldwide. The high number of “unclear” reported (n=53; 15.9%) may be related to the accelerated speed of publication, the forgetfulness of these items in the writing process of the manuscript or the lack of knowledge of checklists like the one used in this work (*13*). Therefore, we recommend the use of standardized checklists for the planning, execution and reporting of prevalence studies. Intermediate risk of bias was associated with: American and European studies, blood samples, P>1%, and internal/external validation. Low risk of bias was associated with: Asian studies, P<1%, NPS samples and RT-PCR tests. Although the number of studies were low and the correspondence analysis presents some outliers due to the low representativeness of some categories, these conclusions were highly consistent and showed some important aspects of population-based COVID-19 prevalence studies associated with the methodological quality.

## Supporting information

Apendix 1 - PROSPERO protocol

Summary table

## Data Availability

All data from this manuscript is accessible upon request to the corresponding author.

## Acknowledgements

The authors would like to thank the included studies for the data provided and for important contributions in facing the pandemic in their countries.

## Disclaimers

The authors declare no competing interests.

## Funding

This work was supported by grants from Coordenação de Aperfeiçoamento de Pessoal de Nível Superior – Brasil (CAPES) – Finance Code 001, Conselho Nacional de Desenvolvimento Científico e Tecnológico (CNPq), and Fundação de Amparo à Pesquisa do Estado do RS (FAPERGS) (Brazilian Government Agencies). The funders had no role in the study design, data generation and analysis, decision to publish or the preparation of the manuscript.

**Table**. Characteristics of 37 population-based prevalence surveys during the COVID-19 pandemic until September.

**Appendix 1**. PROSPERO protocol.

**Appendix 2, Checklist**. PRISMA Checklist

**Appendix 2, Table 1**. Search strategies for different databases.

**Appendix 2, Table 2**. Articles and reasons for exclusion after full-text review.

**Appendix 2, Figure**. Traffic light plot of the domain-level judgements for each individual study.

## Notes

### Competing Interest Statement

The authors have declared no competing interest.

### Clinical Protocols

https://www.crd.york.ac.uk/prospero/display_record.php?ID=CRD42020202186

### Funding Statement

This work was supported by grants from Coordenacao de Aperfeicoamento de Pessoal de Nivel Superior - Brazil (CAPES) - Finance Code 001, Conselho Nacional de Desenvolvimento Cientifico e Tecnologico (CNPq), and Fundacao de Amparo a Pesquisa do Estado do RS (FAPERGS) (Brazilian Government Agencies). The funders had no role in the study design, data generation and analysis, decision to publish or the preparation of the manuscript.

### Author Declarations

Since it is a systematic review article, there was no requirement for submission to an ethics committee.

