## Supplementary material for "Population-based prevalence surveys during the COVID-19 pandemic: a systematic review": Apendix 1 - PROSPERO protocol

### Systematic review

#### 1. * Review title.

Give the title of the review in English

#### 2. Original language title.

For reviews in languages other than English, give the title in the original language. This will be displayed with the English language title.

#### 3. * Anticipated or actual start date.

Give the date the systematic review started or is expected to start.

15/07/2020

#### 4. * Anticipated completion date.

Give the date by which the review is expected to be completed.

17/10/2020

#### 5. * Stage of review at time of this submission.

Tick the boxes to show which review tasks have been started and which have been completed. Update this field each time any amendments are made to a published record.

**Reviews that have started data extraction (at the time of initial submission) are not eligible for inclusion in PROSPERO**. If there is later evidence that incorrect status and/or completion date has been supplied, the published PROSPERO record will be marked as retracted.

This field uses answers to initial screening questions. It cannot be edited until after registration.

The review has not yet started: No

| Review stage | Started | Completed |
|---|---|---|
| Preliminary searches | Yes | Yes |
| Piloting of the study selection process | Yes | Yes |
| Formal screening of search results against eligibility criteria | Yes | Yes |
| Data extraction | Yes | Yes |
| Risk of bias (quality) assessment | Yes | Yes |
| Data analysis | Yes | No |

Provide any other relevant information about the stage of the review here.

#### 6. * Named contact.

The named contact is the guarantor for the accuracy of the information in the register record. This may be any member of the review team.

Claudia Elizabeth Thompson

#### Email salutation (e.g. "Dr Smith" or "Joanne") for correspondence:

Ms Thompson

#### 7. * Named contact email.

Give the electronic email address of the named contact.

#### 8. Named contact address

Give the full institutional/organisational postal address for the named contact.

Rua Sarmento Leite, 245, Centro Histórico, Zip code: 90050-170, Porto Alegre, Rio Grande do Sul

#### 9. Named contact phone number.

Give the telephone number for the named contact, including international dialling code.

+55 (51) 3303-8889

#### 10. * Organisational affiliation of the review.

Full title of the organisational affiliations for this review and website address if available. This field may be completed as 'None' if the review is not affiliated to any organisation.

Universidade Federal de Ciências da Saúde de Porto Alegre

#### Organisation web address:

http://www.ufcspa.edu.br

#### 11. * Review team members and their organisational affiliations.

Give the personal details and the organisational affiliations of each member of the review team. Affiliation refers to groups or organisations to which review team members belong. **NOTE: email and country now MUST be entered for each person, unless you are amending a published record.**

Ms Claudia Elizabeth Thompson. Universidade Federal de Ciências da Saúde de Porto Alegre
Ms Liane Nanci Rotta. Universidade Federal de Ciências da Saúde de Porto Alegre
Ms Andressa Schneiders Santos. Universidade Federal de Ciências da Saúde de Porto Alegre
Ms Andressa Barreto Glaeser. Universidade Federal de Ciências da Saúde de Porto Alegre
Ms Ana Trindade Winck. Universidade Federal de Ciências da Saúde de Porto Alegre
Ms Amanda de Menezes Mayer. Universidade Federal do Rio Grande do Sul
Ms Carem Luana Machado Lessa. Universidade Federal de Ciências da Saúde de Porto Alegre
Ms Júlia Gonçalves Küchle. Universidade Federal de Ciências da Saúde de Porto Alegre
Mr Paulo Ricardo Gazzola Zen. Universidade Federal de Ciências da Saúde de Porto Alegre
Mr Vinícius Bonetti Franceschi. Universidade Federal do Rio Grande do Sul
Ms Janini Cristina Paiz. Universidade Federal do Rio Grande do Sul

Mr Gabriel Dickin Caldana. Universidade Federal de Ciências da Saúde de Porto Alegre
Mr Álvaro Vigo. Universidade Federal do Rio Grande do Sul

#### 12. * Funding sources/sponsors.

Details of the individuals, organizations, groups, companies or other legal entities who have funded or sponsored the review.

This study is being financed in part by the Coordenação de Aperfeiçoamento de Pessoal de Nível Superior - Brasil (CAPES)

#### Grant number(s)

State the funder, grant or award number and the date of award

Finance Code 001

#### 13. * Conflicts of interest.

List actual or perceived conflicts of interest (financial or academic).

None

#### 14. Collaborators.

Give the name and affiliation of any individuals or organisations who are working on the review but who are not listed as review team members. **NOTE: email and country must be completed for each person, unless you are amending a published record.**

#### 15. * Review question.

State the review question(s) clearly and precisely. It may be appropriate to break very broad questions down into a series of related more specific questions. Questions may be framed or refined using PI(E)COS or similar where relevant.

What are the main results regarding COVID-19 prevalence obtained through population-based surveys, their confiability and bias?

#### 16. * Searches.

State the sources that will be searched (e.g. Medline). Give the search dates, and any restrictions (e.g. language or publication date). Do NOT enter the full search strategy (it may be provided as a link or attachment below.)

Literature searches will be conducted in the following electronic databases: MEDLINE (accessed via PubMed), Excerpta Medica dataBASE (EMBASE), bioRxiv and medRxiv.

Terms to be used individually or in combination in PubMed include “Serologic Tests [MeSH and entry terms]”, “Real-Time Polymerase Chain Reaction [MeSH and entry terms]”, “Cross-Sectional Studies [MeSH and entry terms]”, “COVID-19 [MeSH and entry terms]”.

The search strategy will be adapted to each database searched.

The same words and synonyms will be used as text words for searching in the title and abstract fields.

#### 17. URL to search strategy.

Upload a file with your search strategy, or an example of a search strategy for a specific database, (including the keywords) in pdf or word format. In doing so you are consenting to the file being made publicly accessible. Or provide a URL or link to the strategy. Do NOT provide links to your search **results**.

Alternatively, upload your search strategy to CRD in pdf format. Please note that by doing so you are consenting to the file being made publicly accessible.

Do not make this file publicly available until the review is complete

#### 18. * Condition or domain being studied.

Give a short description of the disease, condition or healthcare domain being studied in your systematic review.

In December 2019, a novel coronavirus (SARS-CoV-2) was identified as the causative agent of severe acute respiratory syndrome cases in Wuhan, Hubei province, China.

The illness, named COVID-19, then spread rapidly around the world, and was considered a pandemic in March 11, 2020.

COVID-19 causes varying degrees of illness, ranging from asymptomatic infection to pneumonia, with higher fatality rates in elderly and chronically ill patients.

Up to 01 August 2020, ~18, 000, 000 people around the world had been infected, and ~700, 000 deaths reported.

#### 19. * Participants/population.

Specify the participants or populations being studied in the review. The preferred format includes details of both inclusion and exclusion criteria.

Inclusion criteria: populations in countries, states, counties, regions or municipalities affected by the COVID-19 pandemic.Exclusion criteria: (i) studies with correlation between COVID-19 and other diseases or health determinants, (ii) non-random selection of participants (e. g. convenience sampling), (iii) inclusion of a specific group of participants only (e. g., with comorbidities, pregnant, elderly, healthcare workers, pediatric patients), (iv) non-human samples (e.g. air, sewage).

#### 20. * Intervention(s), exposure(s).

Give full and clear descriptions or definitions of the interventions or the exposures to be reviewed. The preferred format includes details of both inclusion and exclusion criteria.

Population-based prevalence surveys during the COVID-19 pandemic.

Two kinds of tests are available for COVID-19: viral tests and antibody tests:

Viral tests (e.g. RT-PCR) detect current infection.

Antibody tests (e.g. point-of care tests, or ELISA) detect a past infection and the production of antibodies against SARS-CoV-2.

Household population-based studies can help to define the role that subclinical, asymptomatic, and mild infections play in transmission, and to guide evidence-based decisions about the prioritization of non-pharmacological control measures while no vaccine is available.

Moreover, accurate estimates of exposed and susceptible populations and the fatality rates can be estimated based on populational evidence.

#### 21. * Comparator(s)/control.

Where relevant, give details of the alternatives against which the intervention/exposure will be compared (e.g. another intervention or a non-exposed control group). The preferred format includes details of both inclusion and exclusion criteria.

Not applicable.

#### 22. * Types of study to be included.

Give details of the study designs (e.g. RCT) that are eligible for inclusion in the review. The preferred format includes both inclusion and exclusion criteria. If there are no restrictions on the types of study, this should be stated.

Inclusion criteria: any cross-sectional or repeated cross-sectional studies measuring the prevalence of COVID-19 from serological or molecular tests in a population in a country, state, county, region or municipality.

Exclusion criteria: studies that are not cross-sectional in design, case reports, case-control studies, cohort studies, or literature reviews; studies investigating the relationship between COVID-19 and other diseases; studies that have undertaken convenience sampling, not minimally representative of the population; studies that have included a specific group of participants (e.g. susceptible patients, hospitalized patients, pregnant women, and children); and studies involving non-human samples (e.g. air, sewage).

#### 23. Context.

Give summary details of the setting or other relevant characteristics, which help define the inclusion or exclusion criteria.

Cross-sectional and repeated-cross sectional studies that include a representative sample of the population, randomly selected (i.e., individuals of both genders, of any age and race or ethnic background, and with any health condition are included).

#### 24. * Main outcome(s).

Give the pre-specified main (most important) outcomes of the review, including details of how the outcome is defined and measured and when these measurement are made, if these are part of the review inclusion criteria.

The main results regarding the prevalence of SARS-CoV-2 in the population, obtained through population-based surveys.

##### * Measures of effect

Please specify the effect measure(s) for you main outcome(s) e.g. relative risks, odds ratios, risk difference, and/or 'number needed to treat.

Population follow-up time.

#### 25. * Additional outcome(s).

List the pre-specified additional outcomes of the review, with a similar level of detail to that required for main outcomes. Where there are no additional outcomes please state ‘None’ or ‘Not applicable’ as appropriate to the review

The confiability and bias of population-based survey results.

#### * Measures of effect

Please specify the effect measure(s) for you additional outcome(s) e.g. relative risks, odds ratios, risk difference, and/or 'number needed to treat.

Population follow-up time.

#### 26. * Data extraction (selection and coding).

Describe how studies will be selected for inclusion. State what data will be extracted or obtained. State how this will be done and recorded.

The study will be developed by four groups of authors (CS and ATW, AS and VF, AC and JK, JCP and GDC).

Each pair of reviewers will separately and independently screen the titles and abstracts of the studies identified from initial searches, and a standard screening checklist based on the eligibility criteria listed above will be employed for each study.

Studies that do not meet the criteria according to the titles or abstracts will be excluded.

Full text versions of the remaining studies, including those that are potentially eligible, and studies for which eligibility is uncertain, will be retrieved for a second review by at least two independent reviewers to determine inclusion/exclusion.Disagreements with regard to the study eligibility will be further discussed among the reviewers, and if consensus cannot be reached, a third reviewer (ATW) will make the ultimate decision.

If more than one publication reports the results from the same study population, we will choose the publication with the largest sample size.

The four pairs of review authors will then separately and independently extract the following data from the studies selected for inclusion: authors, study location, coverage, study type, random sampling method, period of testing, number of tests, biological samples, type of test used, if test validation was performed, test sensitivity and specificity, prevalence, and statistical methods.

Disagreements regarding data extraction between the authors will be resolved by discussion and consensus, or, if consensus cannot be reached, a third author (ATW) will review the study and arbitrate.

#### 27. * Risk of bias (quality) assessment.

State which characteristics of the studies will be assessed and/or any formal risk of bias/quality assessment tools that will be used.

Reviewers will independently and in pairs assess the risk of bias in the included studies using Joanna Briggs Institute (JBI) Critical Appraisal Checklist for Prevalence Studies.

#### 28. * Strategy for data synthesis.

Describe the methods you plan to use to synthesise data. This **must not be generic text** but should be **specific to your review** and describe how the proposed approach will be applied to your data. If meta-analysis is planned, describe the models to be used, methods to explore statistical heterogeneity, and software package to be used.

Variables will be narratively synthetized and summarized using descriptive statistics. We will also perform a correspondence analysis (CA), which is a multivariate statistical technique conceptually similar to principal component analysis, but applies to categorical rather than continuous data. It provides a means of displaying or summarising a set of data in two-dimensional graphical form. We will use information such as the study location, coverage, study type, random sampling method, biological samples, type of test used, if test validation was performed, test sensitivity and specificity, and prevalence.

#### 29. * Analysis of subgroups or subsets.

State any planned investigation of ‘subgroups’. Be clear and specific about which type of study or participant will be included in each group or covariate investigated. State the planned analytic approach.

If a sufficient number of randomized trials are identified, a subgroup analysis will be performed according tothe type of comparator.

#### 30. * Type and method of review.

Select the type of review, review method and health area from the lists below.

##### Type of review

Cost effectiveness
No

Diagnostic
No

Epidemiologic
Yes

Individual patient data (IPD) meta-analysis
No

Intervention
No

Meta-analysis
No

Methodology
Yes

Narrative synthesis
No

Network meta-analysis
No

Pre-clinical
No

Prevention
No

Prognostic
No

Prospective meta-analysis (PMA)

No

Review of reviews
No

Service delivery
No

Synthesis of qualitative studies
No

Systematic review
Yes

Other
No

#### Health area of the review

Alcohol/substance misuse/abuse
No

Blood and immune system
Yes

Cancer
No

Cardiovascular
No

Care of the elderly
No

Child health
No

Complementary therapies
No

COVID-19
Yes

For COVID-19 registrations please tick all categories that apply. Doing so will enable your record to appear in area-specific searches

Chinese medicine
Diagnosis
Epidemiological
Genetics
Health impacts
Mental health
PPE
Prognosis
Public health
Rehabilitation
Service delivery
Transmission
Treatments
Vaccines
Other

Crime and justice
No

Dental
No

Digestive system
No

Ear, nose and throat
No

Education
No

Endocrine and metabolic disorders
No

Eye disorders
No

General interest
No

Genetics
No

Health inequalities/health equity
No

Infections and infestations
Yes

International development
No

Mental health and behavioural conditions
No

Musculoskeletal
No

Neurological
No

Nursing
No

Obstetrics and gynaecology
No

Oral health
No

Palliative care
No

Perioperative care
No

Physiotherapy
No

Pregnancy and childbirth

No

Public health (including social determinants of health)
Yes

Rehabilitation
No

Respiratory disorders
Yes

Service delivery
No

Skin disorders
No

Social care
Yes

Surgery
No

Tropical Medicine
No

Urological
No

Wounds, injuries and accidents
No

Violence and abuse
No

#### 31. Language.

Select each language individually to add it to the list below, use the bin icon to remove any added in error.
English

There is not an English language summary

#### 32. * Country.

Select the country in which the review is being carried out. For multi-national collaborations select all the countries involved.

Brazil

#### 33. Other registration details.

Name any other organisation where the systematic review title or protocol is registered (e.g. Campbell, or The Joanna Briggs Institute) together with any unique identification number assigned by them. If extracted data will be stored and made available through a repository such as the Systematic Review Data Repository (SRDR), details and a link should be included here. If none, leave blank.

#### 34. Reference and/or URL for published protocol.

If the protocol for this review is published provide details (authors, title and journal details, preferably in Vancouver format)

Add web link to the published protocol.

Or, upload your published protocol here in pdf format. Note that the upload will be publicly accessible.

No I do not make this file publicly available until the review is complete

Please note that the information required in the PROSPERO registration form must be completed in full even if access to a protocol is given.

#### 35. Dissemination plans.

Do you intend to publish the review on completion?

Yes

Give brief details of plans for communicating review findings.?

#### 36. Keywords.

Give words or phrases that best describe the review. Separate keywords with a semicolon or new line. Keywords help PROSPERO users find your review (keywords do not appear in the public record but are included in searches). Be as specific and precise as possible. Avoid acronyms and abbreviations unless these are in wide use.

SARS-CoV-2; COVID-19; Prevalence; Anti-SARS-CoV-2; Epidemiology; Infectivity, Infectious Diseases

#### 37. Details of any existing review of the same topic by the same authors.

If you are registering an update of an existing review give details of the earlier versions and include a full bibliographic reference, if available.

#### 38. * Current review status.

Update review status when the review is completed and when it is published.New registrations must be ongoing.
Please provide anticipated publication date

Review_Ongoing

#### 39. Any additional information.

Provide any other information relevant to the registration of this review.

#### 40. Details of final report/publication(s) or preprints if available.

Leave empty until publication details are available OR you have a link to a preprint. List authors, title and journal details preferably in Vancouver format.

Give the link to the published review or preprint.
