## Appendix 2 for "Population-based prevalence surveys during the COVID-19 pandemic: a systematic review"

### Checklist. PRISMA Checklist

| **Section/topic** | **#** | **Checklist item** | **Reported on section or paragraph** |
| --- | --- | --- | --- |
| **TITLE** | | |  |
| Title | 1 | Identify the report as a systematic review, meta-analysis, or both. | Title |
| **ABSTRACT** | | |  |
| Structured summary | 2 | Provide a structured summary including, as applicable: background; objectives; data sources; study eligibility criteria, participants, and interventions; study appraisal and synthesis methods; results; limitations; conclusions and implications of key findings; systematic review registration number. | Abstract (as possible within journal word limits) |
| **INTRODUCTION** | | |  |
| Rationale | 3 | Describe the rationale for the review in the context of what is already known. | Introduction  (par. 1-4) |
| Objectives | 4 | Provide an explicit statement of questions being addressed with reference to participants, interventions, comparisons, outcomes, and study design (PICOS). | Introduction  (par. 5) |
| **METHODS** | | |  |
| Protocol and registration | 5 | Indicate if a review protocol exists, if and where it can be accessed (e.g., Web address), and, if available, provide registration information including registration number. | Methods  (Registration and Reporting) |
| Eligibility criteria | 6 | Specify study characteristics (e.g., PICOS, length of follow-up) and report characteristics (e.g., years considered, language, publication status) used as criteria for eligibility, giving rationale. | Methods  (Inclusion and Exclusion Criteria) |
| Information sources | 7 | Describe all information sources (e.g., databases with dates of coverage, contact with study authors to identify additional studies) in the search and date last searched. | Methods  (Search Strategy) |
| Search | 8 | Present full electronic search strategy for at least one database, including any limits used, such that it could be repeated. | Appendix 2, Table 1 |
| Study selection | 9 | State the process for selecting studies (i.e., screening, eligibility, included in systematic review, and, if applicable, included in the meta-analysis). | Methods (Article Screening and Data Extraction) |
| Data collection process | 10 | Describe method of data extraction from reports (e.g., piloted forms, independently, in duplicate) and any processes for obtaining and confirming data from investigators. | Methods (Article Screening and Data Extraction) |
| Data items | 11 | List and define all variables for which data were sought (e.g., PICOS, funding sources) and any assumptions and simplifications made. | Methods (Article Screening and Data Extraction;  Definitions; Data analysis) |
| Risk of bias in individual studies | 12 | Describe methods used for assessing risk of bias of individual studies (including specification of whether this was done at the study or outcome level), and how this information is to be used in any data synthesis. | Methods (Survey Quality) |
| Summary measures | 13 | State the principal summary measures (e.g., risk ratio, difference in means). | Results (par 1-3) |
| Synthesis of results | 14 | Describe the methods of handling data and combining results of studies, if done, including measures of consistency (e.g., I^2^) for each meta-analysis. | N/A |
| Risk of bias across studies | 15 | Specify any assessment of risk of bias that may affect the cumulative evidence (e.g., publication bias, selective reporting within studies). | N/A |
| Additional analyses | 16 | Describe methods of additional analyses (e.g., sensitivity or subgroup analyses, meta-regression), if done, indicating which were pre-specified. | Methods (Data analysis) |
| **RESULTS** | | |  |
| Study selection | 17 | Give numbers of studies screened, assessed for eligibility, and included in the review, with reasons for exclusions at each stage, ideally with a flow diagram. | Results (par. 1)  Appendix 2, Table 2 |
| Study characteristics | 18 | For each study, present characteristics for which data were extracted (e.g., study size, PICOS, follow-up period) and provide the citations. | Table |
| Risk of bias within studies | 19 | Present data on risk of bias of each study and, if available, any outcome level assessment (see item 12). | Results (par. 3)  Appendix 2, Figure 1 |
| Results of individual studies | 20 | For all outcomes considered (benefits or harms), present, for each study: (a) simple summary data for each intervention group (b) effect estimates and confidence intervals, ideally with a forest plot. | N/A |
| Synthesis of results | 21 | Present results of each meta-analysis done, including confidence intervals and measures of consistency. | N/A |
| Risk of bias across studies | 22 | Present results of any assessment of risk of bias across studies (see Item 15). | N/A |
| Additional analysis | 23 | Give results of additional analyses, if done (e.g., sensitivity or subgroup analyses, meta-regression [see Item 16]). | Results (par. 4-5) |
| **DISCUSSION** | | |  |
| Summary of evidence | 24 | Summarize the main findings including the strength of evidence for each main outcome; consider their relevance to key groups (e.g., healthcare providers, users, and policy makers). | Discussion  (par. 1-10) |
| Limitations | 25 | Discuss limitations at study and outcome level (e.g., risk of bias), and at review-level (e.g., incomplete retrieval of identified research, reporting bias). | Discussion  (par. 11-12) |
| Conclusions | 26 | Provide a general interpretation of the results in the context of other evidence, and implications for future research. | Discussion  (par. 13) |
| **FUNDING** | | |  |
| Funding | 27 | Describe sources of funding for the systematic review and other support (e.g., supply of data); role of funders for the systematic review. | Funding statement |

### Table 1. Search strategies for different databases

| **Set** | **Pubmed** | **Embase** | **BioRxiv *** | **MedRxiv *** |
| --- | --- | --- | --- | --- |
| 1 | (“Serologic Tests”[Mesh] OR “Serology”[Mesh] OR “Immunoassay”[Mesh] OR “Enzyme-Linked Immunosorbent Assay”[Mesh] OR antibody* OR “ELISA” OR “Lateral Flow Immunoassay” OR LFIA OR “Chemiluminescent Immunoassay” OR CLIA OR IgG OR IgM OR “Real-Time Polymerase Chain Reaction”[Mesh] OR “Reverse Transcriptase Polymerase Chain Reaction”[Mesh] OR “viral PCR” OR “RT-PCR” OR “qRT-PCR” OR “multiplex PCR” OR “targeted testing” OR swab*) | ('serology'/exp OR 'immunoassay'/exp OR 'enzyme linked immunosorbent assay'/exp OR antibody* OR 'lateral flow immunoassay'/exp OR 'chemiluminescent immunoassay'/exp OR 'immunoglobulin g'/exp OR 'immunoglobulin m'/exp OR 'real time polymerase chain reaction'/exp OR 'reverse transcription polymerase chain reaction'/exp OR 'real time reverse transcription polymerase chain reaction'/exp OR 'viral pcr' OR 'rt-pcr' OR 'qrt-pcr' OR 'multiplex polymerase chain reaction'/exp OR 'targeted testing' OR swab*) | (“cross-sectional” OR “population-based” OR seroprevalence OR prevalence) | (“cross-sectional” OR “population-based” OR seroprevalence OR prevalence) |
| 2 | (“Cross-Sectional Studies”[Mesh] OR “population-based” OR “population screening” OR seroprevalence OR “prevalence” OR “survey” OR serosurvey OR serosurveys) | ('cross-sectional study'/exp OR 'population-based' OR 'population screening'/exp OR 'seroprevalence'/exp OR 'prevalence'/exp OR 'survey'/exp OR serosurvey OR serosurveys) | (“SARS-CoV-2” OR “COVID-19”) | (“SARS-CoV-2” OR “COVID-19”) |
| 3 | (“severe acute respiratory syndrome coronavirus 2” OR “2019-nCoV” OR “SARS-CoV-2” OR “COVID-19”) | ('severe acute respiratory syndrome coronavirus 2'/exp ' OR '2019-ncov'/exp OR 'sars-cov-2'/exp OR 'covid-19'/exp) | **Filter**: 01 Dec, 2019 and 31 Oct, 2020 | **Filter**: 01 Dec, 2019 and 31 Oct, 2020 |
| 4 | **Filters**: “English” + “1 year” | [english]/lim AND [embase]/lim) AND 2020:py | 1 AND 2 AND 3 | 1 AND 2 AND 3 |
| 5 | 1 AND 2 AND 3 AND 4 | 1 AND 2 AND 3 AND 4 |  |  |

* Databases with character limits in the search and without controlled vocabulary

### Table 2. Articles and reasons for exclusion after full-text review

| **Title** | **Authors** | **DOI** | **Database** | **Exclusion criteria (EC)** |
| --- | --- | --- | --- | --- |
| Seroprevalence of immunoglobulin M and G antibodies against SARS-CoV-2 in China | Xu et al. | [10.1038/s41591-020-0949-6](http://doi.org/10.1038/s41591-020-0949-6) | Pubmed | EC1, EC3, EC5 |
| SARS-CoV-2 in rural Latin America. A population-based study in coastal Ecuador | Del Brutto et al. | [10.1093/cid/ciaa1055](http://doi.org/10.1093/cid/ciaa1055) | Pubmed | EC5 |
| Detection, prevalence, and duration of humoral responses to SARS-CoV-2 under conditions of limited population exposure | Ripperger et al. | [10.1101/2020.08.14.20174490](http://doi.org/10.1101/2020.08.14.20174490) | Pubmed | EC3 |
| Prevalence of Antibodies to SARS-CoV-2 in Italian Adults and Associated Risk Factors | Vena et al. | [10.1093/cid/ciaa1234](http://doi.org/10.1093/cid/ciaa1234) | Pubmed | EC3 |
| Estimation of seroprevalence of novel coronavirus disease (COVID-19) using preserved serum at an outpatient setting in Kobe, Japan: A cross-sectional study. | Doi et al. | [10.1101/2020.04.26.20079822](http://doi.org/10.1101/2020.04.26.20079822) | medRxiv | EC3 |
| Seroprevalence of antibodies against SARS-CoV-2 among public community and health-care workers in Alzintan City of Libya | Kammon et al. | [10.1101/2020.05.25.20109470](http://doi.org/10.1101/2020.05.25.20109470) | medRxiv | EC3 |
| SARS-CoV-2 IgG Antibody Responses in New York City | Reifer et al. | [10.1101/2020.05.23.20111427](http://doi.org/10.1101/2020.05.23.20111427) | medRxiv | EC3, EC5 |
| SARS-CoV-2 Community Transmission During Shelter-in-Place in San Francisco | Chamie et al. | [10.1101/2020.05.25.20109470](http://doi.org/10.1101/2020.05.25.20109470) | medRxiv | EC3 |
| Community prevalence of SARS-CoV-2 in England: Results from the ONS Coronavirus Infection Survey Pilot | Pouwels et al. | [10.1101/2020.05.23.20111427](http://doi.org/10.1101/2020.05.23.20111427) | medRxiv | EC6 |
| Sero-prevalence findings from metropoles in Pakistan: implications for assessing COVID-19 prevalence and case-fatality within a dense, urban working population | Javed et al. | [10.1101/2020.08.13.20173914](http://doi.org/10.1101/2020.08.13.20173914) | medRxiv | EC3, EC5 |
| Universal PCR and antibody testing demonstrate little to no transmission of SARS-CoV-2 in a rural community | Appa et al. | [10.1101/2020.08.15.20175786](https://doi.org/10.1101/2020.08.15.20175786) | medRxiv | EC3 |
| Serial population based serosurvey of antibodies to SARS-CoV-2 in a low and high transmission area of Karachi, Pakistan | Nisar et al. | [10.1101/2020.07.28.20163451](https://doi.org/10.1101/2020.07.28.20163451) | medRxiv | EC3 |

**Exclusion criteria:**

**EC1.** Non-transversal studies (e. g., case reports, case-control, cohort, evaluation and / or design of diagnostic tests only, literature reviews, etc);

**EC3.** Non-random selection of participants (e. g., people attending a specific place, convenience sample);

**EC5.** Inclusion of a specific group of participants only (e. g., people with comorbidities, pregnant women, more susceptible patients, women, pediatric patients);

**EC6.** Inconsistencies in the data presented regarding sample size.

### Figure. Traffic light plot of the domain-level judgements for each individual study

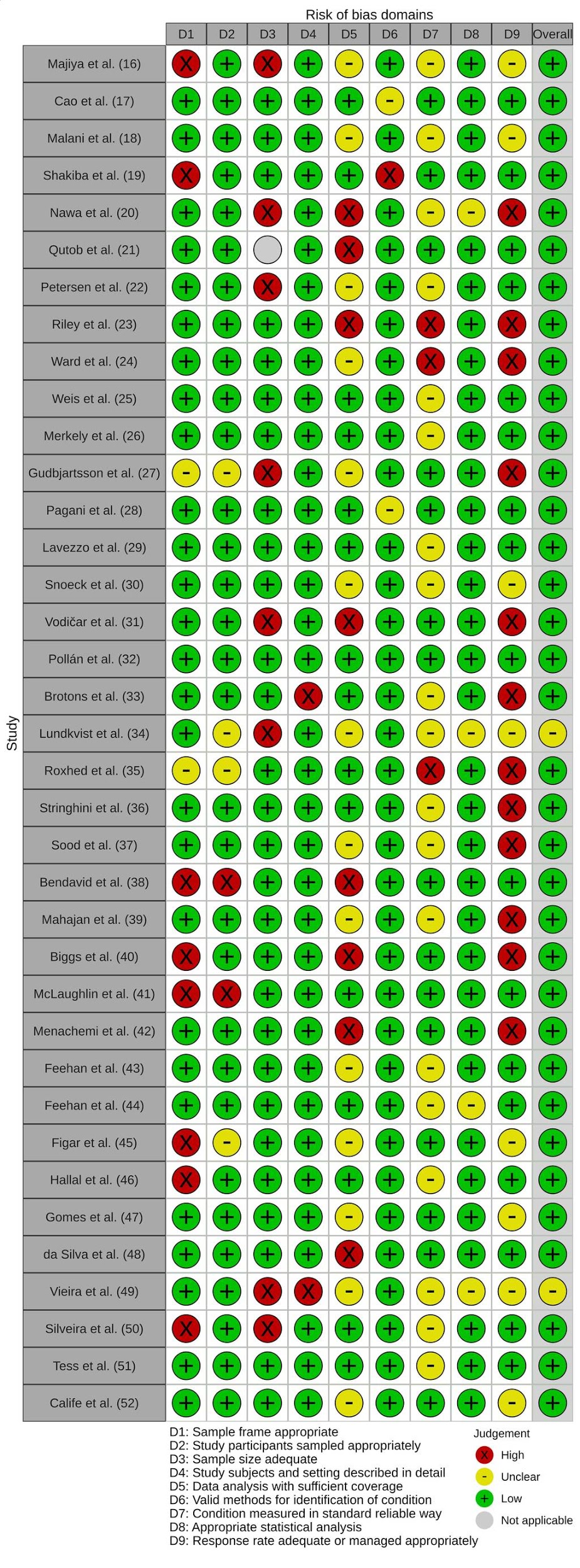

^a^ Overall risk of bias was counted so that the category with the most representativeness (low, unclear or high) in the studies was selected. In the event of a tie, if the sum of unclear and high was greater than low risk of bias, the category with the highest representation between unclear and high was selected.
