## Supplementary material for "Population-based prevalence surveys during the COVID-19 pandemic: a systematic review": Summary table

**Table.** Characteristics of 37 population-based prevalence surveys during the COVID-19 pandemic until September

| **Continent and Region** | **Coverage** | **no. rounds** | **Period (2020)** | **Sample selection method** | **no. tests** | **Biological samples** | **Test(s) used** | **Test validation †** | **Sensitivity**  **(95% CI) ‡** | **Specificity (95% CI) ‡** | **Prevalence (95% CI)** | **Ref** |
| --- | --- | --- | --- | --- | --- | --- | --- | --- | --- | --- | --- | --- |
| **Africa** |  |  |  |  |  |  |  |  |  |  |  |  |
| Niger State / Nigeria | State | 1 | June 26-30 | Random selection using clustered-stratified strategy covering the 3 geopolitical zones in the state | 185 | Whole blood | LFIA for IgG/IgM | Yes, RT-PCR confirmed cases | 100% | 100% | IgG: 25.41%  IgM: 2.16% | ([16](https://doi.org/10.1101/2020.08.04.20168112)) |
| **Asia** |  |  |  |  |  |  |  |  |  |  |  |  |
| Wuhan / Hubei / China | Municipality | 1 | May 14 - June 1 | Screening programme (not detailed) | 9,899,828 | NPS | RT-PCR and mixed testing via sample pooling | NA | - | - | 0.303 / 10,000 (0.270 - 0.339) | ([17](https://doi.org/10.1101/2020.06.29.20142554)) |
| Mumbai / India | Municipality | 1 | Slums: June 29 - July 14  Non-slums: July 3 - July 19 | Random selection of households using geographically-spaced community sampling in areas classified as slums and as non-slums | 6,904  Slums: 4,202  No-slums: 2,702 | Serum | CLIA for IgG | No, by other studies | 90% (74.4-96.5)-  96.9% (89.5-99.5) | 100% (95.4-100) | Slums: 54.1% (52.7-55.6)  Non-slums: 16.1% (14.9-17.4) | ([18](http://doi.org/10.1101/2020.08.27.20182741)) |
| Guilan Province / Iran | Province | 1 | April | Random selection of households using multistage cluster strategy | 528 | Capillary blood | LFIA for IgG/IgM | No, by manufacturer and other study | 63.30% | 100% | Raw: 22% (19-26)  Weighted: 21% (14-29)  Test adjusted: 33% (28-39) | ([19](https://doi.org/10.1101/2020.04.26.20079244)) |
| Utsunomiya / Japan | Municipality | 1 | 14 June - 5 July | Random selection of households from basic resident registry | 742 | Uninformed | CLIA for IgG | No, by manufacturer | 97.30% | 96.30% | Unweighted: 0.40% (0.08-1.18)  Weighted: 1.23% (0.17-2.28%) | ([20](https://doi.org/10.1101/2020.07.20.20155945)) |
| West Bank / Palestina | County | 1 | June 15-30 | Random selection of households based on census tracts with probability proportional to size sampling | 1,319 | Serum | CLIA for IgG/IgM | Yes, RT-PCR confirmed cases | 100% (88.1-100) (14 days post PCR confirmation) | 99.81% (99.65-99.9) | 0% (0-0.0036) | ([21](http://doi.org/10.1101/2020.08.28.20180083)) |
| **Europe** |  |  |  |  |  |  |  |  |  |  |  |  |
| Faroe Islands / Denmark | Region | 1 | April 27 - May 1 | Random selection based on the Population Registry | 1,075 | Serum | Automated (ELISA) | No, by manufacturer | 94.4% (90.9-96.8) | 100% (98.8–100.0) | 0.7% (0.3-1.3) | ([22](http://doi.org/10.3201/eid2611.202736)) |
| England | Country | 1 | May 01 - June 01 | Random selection using the National Health Service (NHS) patient list | 120,620 | Self-administered NPS | RT-PCR | NA | 70% | - | 0.13% (0.11-0.15) | ([23](https://doi.org/10.1101/2020.07.10.20150524)) |
| England | Country | 1 | June 20 - 13 July | Random selection of adults using the NHS patient list | 105,651 | Finger-prick blood | LFIA | Yes, RT-PCR confirmed cases | 84.4% (70.5-93.5) | 98.6% (97.1-99.4) | Raw: 5.6% (5.4-5.7).  Test adjusted: 6.0% (5.8-6.1) | ([24](https://doi.org/10.1101/2020.08.12.20173690)) |
| Neustadt-am-Rennsteig / Germany | Municipality | 1 | May 12-22 | All households from the community were invited | 620 | Pharyngeal washes  Blood drawn | RT-PCR, 2 ELISAs, 2 CLIAs for IgG, 1 CMIA for IgG | Uninformed | - | - | RT-PCR: 0%  Antibody: 8.4% | ([25](https://doi.org/10.1101/2020.07.15.20154112)) |
| Hungary | Country | 1 | May 1-16 | Random selection based on settlements using two-stage stratified strategy, and stratification by age from the population registry | 10,575 | NPS  Blood | RT-PCR  Automated antibody test | Uninformed | - | - | 68 / 10.000 (50–86) | ([26](http://doi.org/10.1007/s11357-020-00226-9)) |
| Iceland | Country | 1 | April 1-4 | Random selection (not detailed) | 2,283 | NPS | RT-PCR | NA | 6 genome copies per reaction | No cross-reactivity observed | 0.6% (0.3-0.9) | ([27](http://doi.org/10.1056/NEJMoa2006100)) |
| Castiglione d'Adda / Lombardy / Italy | Municipality | 1 | May 18 - June 7 | Random selection stratifying by sex and age classes from the municipal registry list | 509 | NPS  Serum blood drawn | CLIA for IgG | Uninformed | - | - | 22.6% (17.2-29.1%) | ([28](https://doi.org/10.1101/2020.06.24.20138875)) |
| Vó / Vêneto / Italy | Municipality | 2 | Beginning of lockdown: February 21–29,  End of lockdown: March 7 | Sampling from the majority of the municipality population | Beginning of lockdown: 2,812  End of lockdown: 2,343 * | NPS | RT-PCR | NA | E gene: 5 genome copies per reaction  RdRp gene: 50 genome copies per reaction | - | Beginning of lockdown: 2.6% (2.1-3.3)  End of lockdown: 1.2% (0.8-1.8) | ([29](http://doi.org/10.1038/s41586-020-2488-1)) |
| Luxembourg | Country | 1 | April 15 - May 5 | Random selection defined by the crossing of the 3 stratification variables through a deterministic random bit generator within strata | RT-PCR: 1,842  IgA and IgG: 1,820 | NPS  Blood drawn | RT-PCR  CE-labelled ELISA for IgA/IgG | Yes, cohort of hospitalized patients | Combined (IgA/IgG): 85.7% | Combined (IgA/IgG) 99,5% | RT-PCR: 0.3%  IgA: 11.0%  IgG: 1.9%  Both IgA and IgG: 1.6% | ([30](https://doi.org/10.1101/2020.05.11.20092916)) |
| Slovenia | Country | 1 | April 20 - May 1 | Random selection of a representative sample using Central Population Register data | 1,366 | NPS | Two-target PCR-based assay | Yes | 100% | 100% | 0.15% (posterior mean 0.18%, 95% Bayesian CI: 0.03-0.47; 95% highest density region 0.01-0.41) | ([31](http://doi.org/10.1016/j.cmi.2020.07.013)) |
| Spain | Country | 1 | April 27 - May 11 | Random selection of households based on census tracts using stratified two-stage strategy and performed by National Institute of Statistics | 61,075 | Finger-prick blood  Blood drawn | LFIA for IgG/IgM  CLIA for IgG | Yes, RT-PCR-positive individuals for both tests | IgG: 82.1%  IgM: 69.6% | IgG: 100%  IgM: 99% | Point-of-care test: 5.0% (4.7-5.4)  Immunoassay: 4.6% (4.3-5.0) | ([32](http://doi.org/10.1016/S0140-6736(20)31483-5)) |
| Barcelona / Spain | Municipality | 1 | April 21-24 | Random selection from individuals registered at a primary health care facility | 311 | Capillary blood | LFIA for IgG/ IgM | Uninformed | - | - | 5.47% (3.44-8.58) | ([33](https://doi.org/10.1101/2020.06.13.20130575)) |
| Stockholm / Sweden | Municipality | 1 | June 17-18 | Random selection (not detailed) | 213  Norra Djurgårdsstaden: 123  Tensta: 90 | Uninformed | LFIA for IgG/IgM | Yes, serum from negative and RT-PCR-positive individuals | ≅100% | 100-95.5% | Norra Djurgårdsstaden: 4.1% (± 3.5%)  Tensta: 30% (± 9.7%) | ([34](http://doi.org/10.1080/20008686.2020.1806505)) |
| Stockholm / Sweden | Municipality | 2 | April 01 - May 31 | Random selection of adults in households and mail distribution of home-sampling kits | 878  Set 1: 435  Set 2: 443 | Finger-prick blood | Multiplexed serology assay (developed in this paper) | Yes, compared to ELISA assays (EuroImmun AG) against the S1 and N proteins | 100% | 96-100% (depending on the antigen used) | Set 1: 10.11% (7.31-12.92)  Set 2: 10.84% (7.94-13.73) | ([35](https://doi.org/10.1101/2020.07.01.20143966)) |
| Geneva / Switzerland | Municipality | 5 | April 6 - May 9  (once every week for 5 weeks) | Random selection based on an already existing representative sample of the general population (Bus Santé study) | 2,766 | Peripheral venous blood | Automated (ELISA) | Yes, sera from pre-pandemic negative controls and RT-PCR-positive individuals | 93% | 100% | R1: 4.8% (2.4–8.0)  R2: 8.5% (5.9–11.4)  R3: 10.9% (7.9–14.4)  R4: 6.6% (4.3–9.4)  R5: 10.8% (8.2–13.9) | ([36](http://doi.org/10.1016/S0140-6736(20)31304-0)) |
| **North America** |  |  |  |  |  |  |  |  |  |  |  |  |
| Los Angeles / California / USA | County | 1 | April 10-14 | Random selection with stratification in subgroups based on age, sex, race, and ethnicity distribution | 863 | Uninformed | LFIA | Yes, RT-PCR-positive individuals | 82.7% (76-88.4) | 99.5% (99.2 - 99.7) | 4.06% (exact binomial: 2.84-5.60) | ([37](http://doi.org/10.1001/jama.2020.8279)) |
| Santa Clara / California / USA | County | 1 | April 3-4 | Facebook ads targeting a sample of individuals living within the county by demographic and geographic characteristics and stratification | 3,330 | Capillary blood | LFIA | Yes, RT-PCR-positive individuals | 82.8% (76.0-88.4) | 99.5% (99.2-99.7) | Raw: 1.5% (exact binomial: 1.1-2.0)  Test adjusted: 1.2% (0.7-1.8)  Census-weighted: 2.8% (1.3-4.7) | ([38](https://doi.org/10.1101/2020.04.14.20062463)) |
| Connecticut / USA | State | 1 | June 10 - July 6 | Random selection from landline and cell phone numbers and re-stratification by census designations | 505 | Serum | Automated Immunodiagnostic System | Yes, RT-PCR-positive individuals | 94% (81-99) | - | 3.1% (90% CI: 1.4-4.8) | ([39](https://doi.org/10.1101/2020.08.04.20168203)) |
| DeKalb and Fulton counties / Georgia / USA | County | 1 | April 28 - May 3 | Random selection of households based on two-stage cluster strategy | 696 | Plasma | Automated Immunodiagnostic System | No, by CDC testing laboratory | 93.2% | 99% | 2.5% (1.4-4.5) | ([40](http://doi.org/10.15585/mmwr.mm6929e2)) |
| Blaine / Idaho / USA | County | 1 | May 4-19 | Random selection of volunteers after stratification by ZIP Code, age and gender within ZIP Code | 917 | Plasma | CLIA for IgG | No, by other studies | 92.9-100% (14 days after symptom onset) | 99.6-100% | 22.7% (20.1-25.5) | ([41](http://doi.org/10.1101/2020.07.19.20157198)) |
| Indiana / USA | State | 1 | April 25-29 | Random selection based on a list of residents derived from tax returns, and stratification using public health preparedness districts as sampling strata | 3,658 | NPS  Peripheral venous blood | RT-PCR  CLIA for IgG | No, by manufacturer | 100% (14 days after symptom onset) | 99.6% (14 days after symptom onset) | RT-PCR raw: 1.74% (1.10-2.54)  Antibody raw: 1.01% (0.76–1.45)  Overall estimate: 2.79% (2.02–3.70) | ([42](http://doi.org/10.15585/mmwr.mm6929e1)) |
| Baton Rouge / Louisiana / USA | Region | 1 | July 15-31 | Random selection using a method developed by Public Democracy, choosing between residents with digital ads for recruitment, and re-stratification of volunteers by census designations | 2,138 | NPS  Blood drawn | RT-PCR  Automated (Qualitative immunoglobulin for IgG) | Uninformed | - | - | 6.6% | ([43](http://doi.org/10.1101/2020.08.26.20180968)) |
| Orleans and Jeferson Parishes / Louisiana / USA | County | 1 | May 9-15 | Random selection based on a novel 2-step system developed by Public Democracy considering >50 characteristics, including social determinants of health and Census population data | 2,640 | NPS  Blood drawn | RT-PCR  Automated (Qualitative immunoglobulin for IgG) | No, by CDC and other studies | 100% (95.1-100) (17 days after symptom onset) (Bryan et al., 2020) | 99.90% (Bryan et al., 2020) | Raw: 6.9% (6.0-8.0%)  Census-weighted: 7.8% (7.8-7.9%) | ([44](http://doi.org/10.3201/eid2611.203029)) |
| **South America** |  |  |  |  |  |  |  |  |  |  |  |  |
| Barrio Mugica / Buenos Aires City / Argentina | Municipality | 1 | June 10 - July 1 | Random selection using two-stage strategy using geographical areas determined by the Department of Statistic and Census | 873 | Finger-prick blood | Automated (ELISA) | Yes, RT-PCR confirmed cases | 95% (after 21 days of symptom onset) | 100% | Weighted IgG: 53.4% (52.8-54.1) | ([45](https://doi.org/10.1101/2020.07.14.20153858)) |
| Brazil | Country | 1 | May 14-21 | Random selection of households based on census tracts from sentinel cities in all Brazilian states | 24,995 | Finger-prick blood | LFIA for IgG/IgM | No, pooled estimate based on four validation studies | 84.8% (95% CI: 81.4-87.8) | 99% (95% CI: 97.8-99.7) | 1.4% (1.3-1.6) | ([46](http://doi.org/10.1101/2020.05.30.20117531)) |
| Espírito Santo / Brazil | State | 1 | May 13-15 | Random selection based on census tracts using most populous municipalities in the state and lesser populous municipalities | 5,775  Prevalence step: 4,612  Extension step: 1,163 | Finger-prick blood | LFIA for IgG/IgM | No, by manufacturer | 86.4% | 97.63% | Prevalence step: 2.1% (1.67-2.52)  Extension step: 0.26% (0.05-0.75) | ([47](https://doi.org/10.1101/2020.06.13.20130559)) |
| Maranhão / Brazil | State | 1 | 27 July - 8 August | Random selection of households based on census tracts in three stratified stages in four regions | 3,156 | Serum | Automated CLIA for IgG/IgM | No, by other studies | - | 99.7% | 40.4% (35.6-45.3) | ([48](http://doi.org/10.1101/2020.08.28.20180463)) |
| Teresina / Piauí / Brazil | Municipality | 7 | April 19 - May 31  (1 week interval) | Random selection of households based on the registry of 78 basic health units and stratification by sex, age, and geographical distribution | 6,300 | Uninformed | LFIA for IgG/IgM | No, by manufacturer | 86% | 99% | R1: 0.56% (0.18-1.3)  R2: 0.89% (0.39-1.75)  R3: 1.44% (0.77-2.45)  R4: 2% (1.19-3.14)  R5: 3.78% (2.63-5.24)  R6: 5.78% (4.3-7.3)  R7: 8.33% (6.61–10.33) | ([49](http://doi.org/10.1590/0037-8682-0351-2020)) |
| Rio Grande do Sul / Brazil | State | 3 | April 11 - May 11  (2 week interval) | Random selection of households based on census tracts using multi-stage sampling strategy in sentinel cities | 13,111  R1: 4,151  R2: 4,460  R3: 4,500 | Finger-prick blood | LFIA for IgG/IgM | Yes, RT-PCR confirmed cases | 84.8% (81.4-87.8%) - Pooled  86.4% (82.4-89.6%) - Manufacturer | 99.0% (97.8–99.7%) - Pooled | R1: 0.048% (0.006–0.174)  R2: 0.135% (0.049–0.293)  R3: 0.222% (0.107–0.408) | ([50](http://doi.org/10.1038/s41591-020-0992-3)) |
| São Paulo / Brazil | Municipality | 1 | May 4-12 | Random selection of households in six districts | 517  Randomly-selected: 299  Cohabitants: 218 | Serum blood drawn | CLIA for IgG/IgM | No, by other studies | IgM: 100%  IgG: 100% (20 days after symptom onset) | IgM: 94.1%  IgG: 99.5% (20 days after symptoms onset) | Census-weighted: 4.7% (3.0-6.6) | ([51](https://doi.org/10.1101/2020.06.29.20142331)) |
| Baixada Santista / São Paulo / Brazil | Region | 1 | Uninformed | Random selection of households based on census tracts and stratification by age, gender and living conditions | 2,342 | Uninformed | LFIA for IgG/IgM | Yes, RT-PCR confirmed cases after more  than 14 days of symptoms | - | - | 1.4% (0.93-1.93) | ([52](http://doi.org/10.1101/2020.08.28.20184010)) |

NPS: Nasopharyngeal and oropharyngeal swabs; R: Round; CLIA: Chemiluminescent microparticle immunoassay; LFIA: Lateral flow immunoassay; ELISA: Enzyme-linked immunosorbent assay.

* Repeated people between the first and second rounds.

† If test validation was performed internally by the study or in a publication performed by the same authors. NA was considered when the RT-PCR method (gold standard) was applied.

‡ Sensitivity and specificity reported by the study or the reference cited, according to the “Test validation” column.
